# Quantifying Fit-for-Purpose in Real World Data: Data Grading and FitQ Scores

**DOI:** 10.1101/2024.02.02.24302239

**Authors:** Michael L. Jackson, Raj Manickam, Dan Derieg, Saurabh Gombar, Yen S Low

**Affiliations:** Atropos Health

## Abstract

Real-world evidence (RWE), derived from analysis of RWD, is increasingly used to guide decisions in drug development, regulatory oversight, and clinical decision-making. Evaluating the fitness-for-purpose of RWD sources is one key component to generating transparent RWE. Here, we demonstrate tools that fill two gaps in the data grading literature. These are the need for quantitative data grading scores, and the need for scoring mechanisms that can be run in automated fashion and at scale. The Real World Data Score (RWDS) rates the overall quality and completeness of a RWD source across a range of customizable metrics. The Fitness Quotient (FitQ) grades how well a specific data source fits a specific RWE query. In concert, these tools give producers and consumers of RWE evidence to assess the quality of the underlying RWD.

## Background

Real-world evidence (RWE) in healthcare is of increasing importance for drug development, regulatory oversight, and clinical decision-making.^1–3^ RWE is derived from analysis of real-world data (RWD), which are generated through routine healthcare encounters, wearable devices, disease registries, and other sources.^4^ Given the well-documented challenges for making causal inference based on RWE, numerous stakeholders have discussed best practices for transparent, reproducible, and valid RWE studies.^1,5–7^ These include proposals for characterizing the reliability and relevance of potential RWD sources with respect to a specific RWE evaluation - whether the RWD are “fit for purpose”.^7–11^

While various stakeholders have discussed the importance of identifying fit for purpose RWD, two gaps in the field are evident. The first is the need for quantitative RWD evaluation metrics. The existing literature provides a variety of frameworks and checklists for characterizing the fitness of RWD for use in RWE (e.g. ^7,10–12^). For the most part, these frameworks operate at a high level, describing general steps such as “list key data elements needed for analysis” or “conduct validation analyses to ensure data reliability.” Largely missing are methods for operationalizing these steps to make quantitative data fitness measures. One step in this direction was proposed by Gatto et al.^5^ In a worked example of their Structured Process to Identify Fit-for-Purpose data (SPIFD), they provide Likert scale grading of RWD sources on metrics such as study population and length of patient follow-up. The Likert scales reflect the number of requirements met within various categories but do not give quantitative comparisons of target metrics between data sources. Similarly, the European Medicine Agency used a set of binary or three-point scales to grade multiple data sources but did not otherwise give comparison metrics.^6^

The second gap is the need to extend fitness-for-purpose assessments to the new paradigm of modern multi-site collaboratives, such as the Food and Drug Administration’s (FDA’s) Sentinel Initiative^13^ or the Observational Health Data Science Initiative (OHDSI)^14^. Current data grading frameworks primarily operate in the setting of bespoke RWE analyses. In these bespoke analyses the RWD assessment goal is to identify the (single) best available data source for one specific RWE question. In contrast, collaboratives aim to run large numbers of RWE analyses across as many data sources as possible.^15^ For these networks, evaluating whether a particular RWD source is fit for a specific analysis needs to happen rapidly and at scale, which requires automation. Automated comparisons of RWD content against prespecified targets (e.g. sample size in each study arm) could provide “go/no-go” decisions of whether to run a particular RWE analysis against a specific RWD source. Beyond a binary decision, a quantitative fitness for purpose score could be used to guide the interpretation of estimates from multiple data sources or could serve as a weighting mechanism for combining estimates across RWD sources.

We have developed two RWD scoring metrics which address these gaps: Real World Data Score (RWDS) and Fitness Quotient (FitQ). RWDS is a quantitative score that provides an overall characterization of the size and richness of an RWD source. FitQ is a question-specific quantitative score that grades a RWD source against target metrics for a particular RWE question. FitQ includes measurements in domains such as effective sample size and depth and breadth of data among the subset of the patient population that could contribute to the RWE analysis. In combination, these scores can provide an overall view of the relative size and richness of different RWD sources and can evaluate the fitness of RWD sources for specific questions, using quantitative scores that can be computed automatically and at scale.

In the sections that follow we first describe how we have operationally defined RWDS and FitQ. We then provide examples of the implementation and use of RWDS and FitQ on several RWD sources and RWE analyses. We close with a discussion of limitations, areas for future research, and some broader applications of the scores.

## Defining RWDS for a RWD source

RWDS helps users understand the overall depth and quality of a RWD source and compare multiple RWD sources against each other. We begin by defining five categories of data quality. For each category we develop a series of individual metrics. We emphasize that the specific categories or metrics we have chosen are not the only possible choices; the implementation of RWDS can be customized to other settings.

The main categories we use are:

- Volume: Total patient count;
- Continuity: Duration of clinical history available for patients;
- Recency: Time from record availability to the present;
- Depth: Availability of patient data from various sources such as procedures, prescriptions, or laboratory testing;
- Breadth: Presence of patients with relevant comorbidities or prescriptions.

Within each of these categories, we then define a series of specific metrics (Figure 1). For example, the Continuity category includes metrics such as “proportion of patients with at least 5 years between their first and last recorded event”, while the Recency category includes metrics such as “proportion of patients with at least two events in or after 2021”. Each of these metrics is defined numerically, either as a proportion of patients in the RWD source or as an absolute number of patients. Metrics that are counts (such as total patient counts) are rescaled logarithmically to reduce variance, while proportions are left on the original scale.

**Figure 1:**
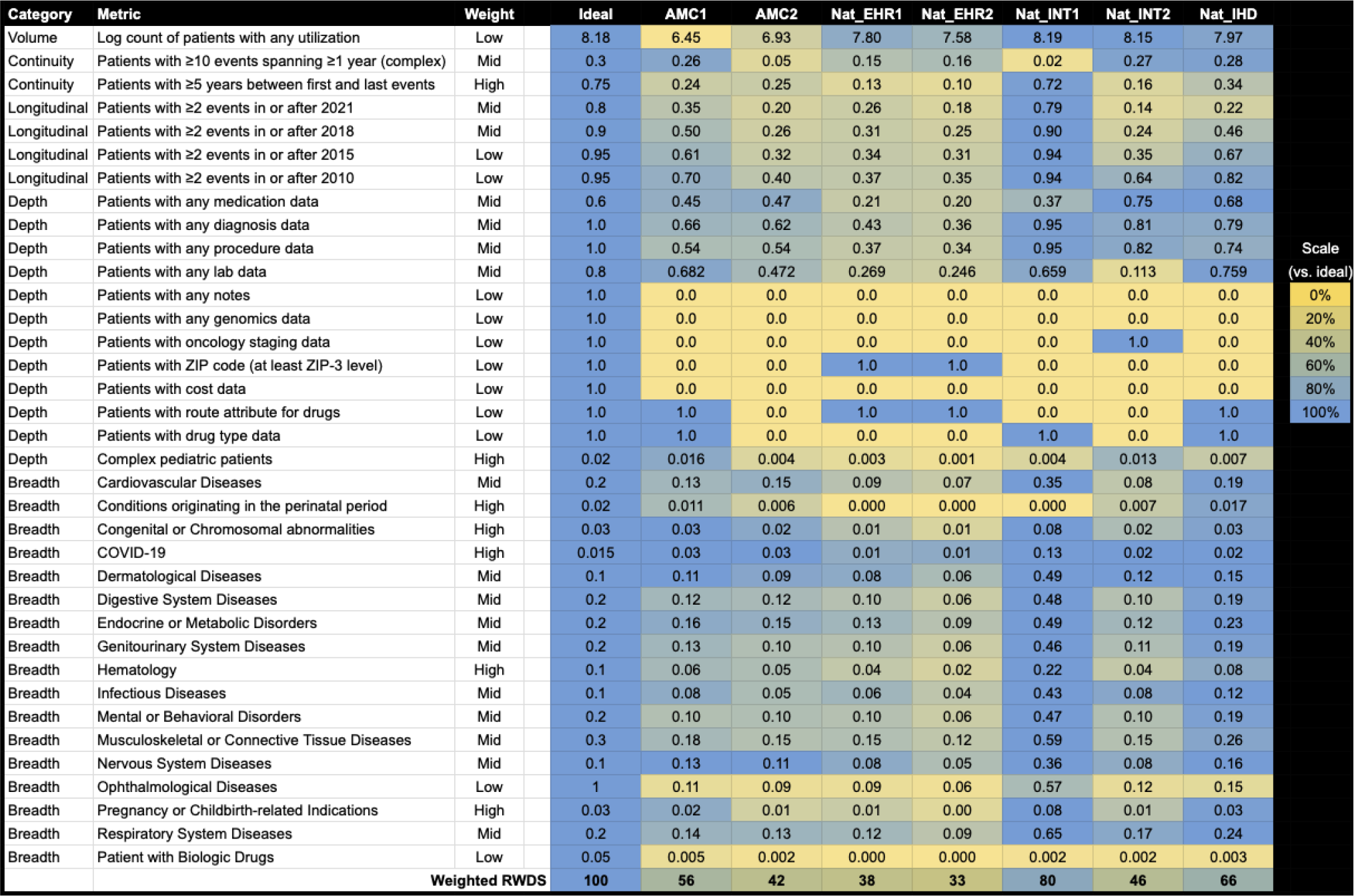
Real World Data Score (RWDS) metrics for six data sources

We then apply weights to each of the metrics to reflect their relative importance in grading the overall volume and completeness of the data. These weights are based on expert determination; the particular values can be adjusted based on contextual needs. For each metric we also describe target value that we might find in a hypothetical ideal RWD source. These targets are used to normalize the weighted scores to a scale from 0 to 100 where 100 represents the hypothetical ideal RWD source of 150 million patients.

A strength of this two-part approach, where weights are applied to calculated metrics, is that the RWDS can be quickly customized for different use cases. Once the individual metrics have been computed, their relative contribution to the RWDS can be modified via the weights. As an example, for a cardiovascular-focused question, the weight for “percent of patients with diagnoses for cardiovascular diseases” can be increased and weights for other chronic conditions can be downgraded.

In addition, optional metrics can be added to RWDS in the event that specific data types are needed or would be beneficial. For example, the presence or frequency of genomics or cancer staging data could be used to evaluate data sources for use in oncology questions.

## Defining FitQ of a RWE question from a RWD source

RWDS as an overall metric of the breadth and depth of a RWD source is complemented by FitQ, an assessment of fitness-for-purpose of a RWD source for a specific RWE question. FitQ begins with a series of metrics which are focused on the actual study population that would be identified from a RWD source (Table 1). The metrics include items related to sample size such as “number of patients in each intervention arm” and items related to depth and breadth of data on these specific subjects, such as “number of days of history prior to index date”.

**Table 1:** FitQ metrics and point values.

| Score components |  | Threshold values |  |  | Points awarded |  |  |
| --- | --- | --- | --- | --- | --- | --- | --- |
| Metric | Description | Best | Good | Fair | Best | Good | Fair |
| Cohort size | Number of patients per intervention arm | 1000 | 250 | 0 | 9 | 6 | 3 |
| Mean days of history | Mean number of days of history prior to index date | 10000 | 2000 | 0 | 3 | 2 | 1 |
| Mean days of followup | Mean number of days of follow-up post index date | 3300 | 1100 | 0 | 3 | 2 | 1 |
| Mean number of medications | Mean number of drug records prior to index date | 45 | 2.5 | 0 | 3 | 2 | 1 |
| Mean number of procedures | Mean number of CPT codes prior to index date | 7 | 3 | 0 | 3 | 2 | 1 |
| Mean number of encounters at | Mean number of encounters (unique dates of features) prior to index date | 4.5 | 1.5 | 0 | 3 | 2 | 1 |
| baseline |  |  |  |  |  |  |  |
| Mean number of diagnostic codes | Mean of sum of number of ICD9-CM and ICD10-CM diagnostic codes prior to index date | 30 | 5 | 0 | 3 | 2 | 1 |
| Comorbidities | Mean of count of number of components of Charlson Comorbidity Index which are zero | 0 | 4 | 8 | 3 | 2 | 1 |
| Outcomes | Mean of proportion(s) of patients with outcome(s) | 0.05 | 0.01 | 0 | 9 | 6 | 3 |
| Overall Grade | Percentage of possible total score (nearest integer). Minimum 33, Maximum: 100 |  |  |  |  |  |  |

For each of these metrics we define threshold values for what would qualify an analytic dataset as “best”, “good”, or “fair” within each category. For example, having at least 1,000 patients in each arm would qualify as “best”, having at least 250 would qualify as “good”, and having fewer than 250 would qualify as “fair”. We then assign point values to each category - typically 3 for best, 2 for good, and 1 for fair, although very important metrics such as cohort size receive more points as a weighting mechanism. The FitQ score is then calculated as the sum of all the point values, normalized so that the maximum possible points would have a FitQ score of 100. As with the RWDS, the relative points assigned to each metric can be customized to provide more or less influence to metrics of interest for a particular RWE question.

## Illustrative example: RWDS

To demonstrate an application of the RWDS, we compute scores for six separate RWD sources. These include:

- AMC1: A large academic medical center with affiliate care sites which provides structured medication, laboratory, procedure, and diagnosis data, with a population of roughly 2.8 million patients. Death data are available from the social security death index;
- AMC2: A large academic medical center with affiliate care sites, which provides structured medication, laboratory, procedure, and diagnosis data, with a population of roughly 8.5 million patients. Death data are available only for inpatient deaths;;
- Nat_IHD: Consolidated data extracted from electronic health records (EHRs) from integrated healthcare delivery (IHD) networks in the United States, comprising medication, laboratory, procedure, diagnosis, and vital sign data, with a population of roughly 107 million patients;
- Nat_EHR1: Consolidated data extracted from EHRs from outpatient and inpatient providers in the United States, including structured medication, laboratory, procedure, and diagnosis data, with a population of roughly 63 million patients. Death data are available but the source is not disclosed to users;
- Nat_EHR2: Consolidated data extracted from EHRs from outpatient and inpatient providers in the United States, including structured medication, laboratory, procedure, and diagnosis data, with a population of roughly 38 million patients;
- Nat_INT1: Integrated claims and EHR records from the United States, covering roughly 153 million patients;
- Nat_INT2: Integrated claims and EHR records from the United States, covering roughly 143 million patients;

Note that while these data come from distinct providers, the patient populations may overlap between data sources.

RWDS scores for these RWD sources range from 33 (Nat_EHR2) to 80 (Nat_INT1). As shown from the individual metrics (Figure 1), the data sources are heterogeneous with respect to depth, breadth, and recency of data.

A few features of the RWDS can be seen from this example. First, the individual metrics themselves can be informative for understanding the relative strengths and limitations of each data source. Second, data sources which have limitations in some areas can still have moderate RWDS scores due to strengths in other areas. For example, AMC1 has a small patient volume (2.8 million patients) compared to the hypothetical ideal (150 million). But a relatively high proportion of patients with longitudinal data and with varying diagnoses compensates to result in a RWDS score of 56. In contrast, Nat_EHR2 has a much larger patient volume (38 million patients), but has lower proportions for metrics related to depth and breadth, resulting in a RWDS of 33.

## Illustrative example: FitQ

Atropos Health provides a RWE consulting service for clinicians, life science researchers, and others.^16^ Users submit questions through Atropos Health’s web portal, where a medical informaticist helps structure the question in Population-Intervention-Control-Outcome-Timeframe (PICOT) format. The requestor can select one or more RWD sources to use for their question, based in part on the RWDS metrics for the data sources. The Atropos medical informaticist translates the question into temporal query language (TQL) code, which extracts patient cohorts from RWD sources.^17^ The extracted data are run through an automated analytic pipeline, and structured results are returned as a report to the medical informaticist, who adds a plain text summary and returns the results to the requestor.

Within this context, the Atropos Health analytic pipeline computes a FitQ score for every RWE analysis. In addition to standard methods for evaluating RWE results (such as sample sizes, p-values, and E-values), these FitQ scores give requestors additional information for making decisions based on the reported findings. For example, a RWE comparison might be the risk of diabetic retinopathy among diabetics treated with rosiglitazone vs. diabetics not treated with rosiglitazone. Analyzing this question using AMC1 data might result in a FitQ of 87, for while the cohort size might only be graded as “fair”, the completeness of other data elements and the frequency of outcomes might all be graded as “best” or “good”.

Here, we summarize FitQ scores for the first 865 consults run on the AMC1, AMC2, and Nat_EHR1 data sources, each of which have over 50 completed consults with FitQ scores. Across all requests, the mean (standard deviation [sd]) FitQ score was 69.2 (sd 11.6) (Figure 2). FitQ scores tended to be highest for consultations using the AMC2 data (mean 73.7, sd 9.7) and lowest for consultations using the Nat_EHR1 data (mean 65.4, sd 11.0). Although Nat_EHR1 was the largest dataset, it tended to score lower on several key metrics of the FitQ score. For example, AMC1 consults scoring better than Nat_EHR1 on the metrics for patient history (mean 2.0 vs. 1.8), patient follow-up (mean 1.7 vs. 1.5), prescriptions (2.8 vs. 2.5), and outcome (mean 6.6 vs 5.6).

**Figure 2:**
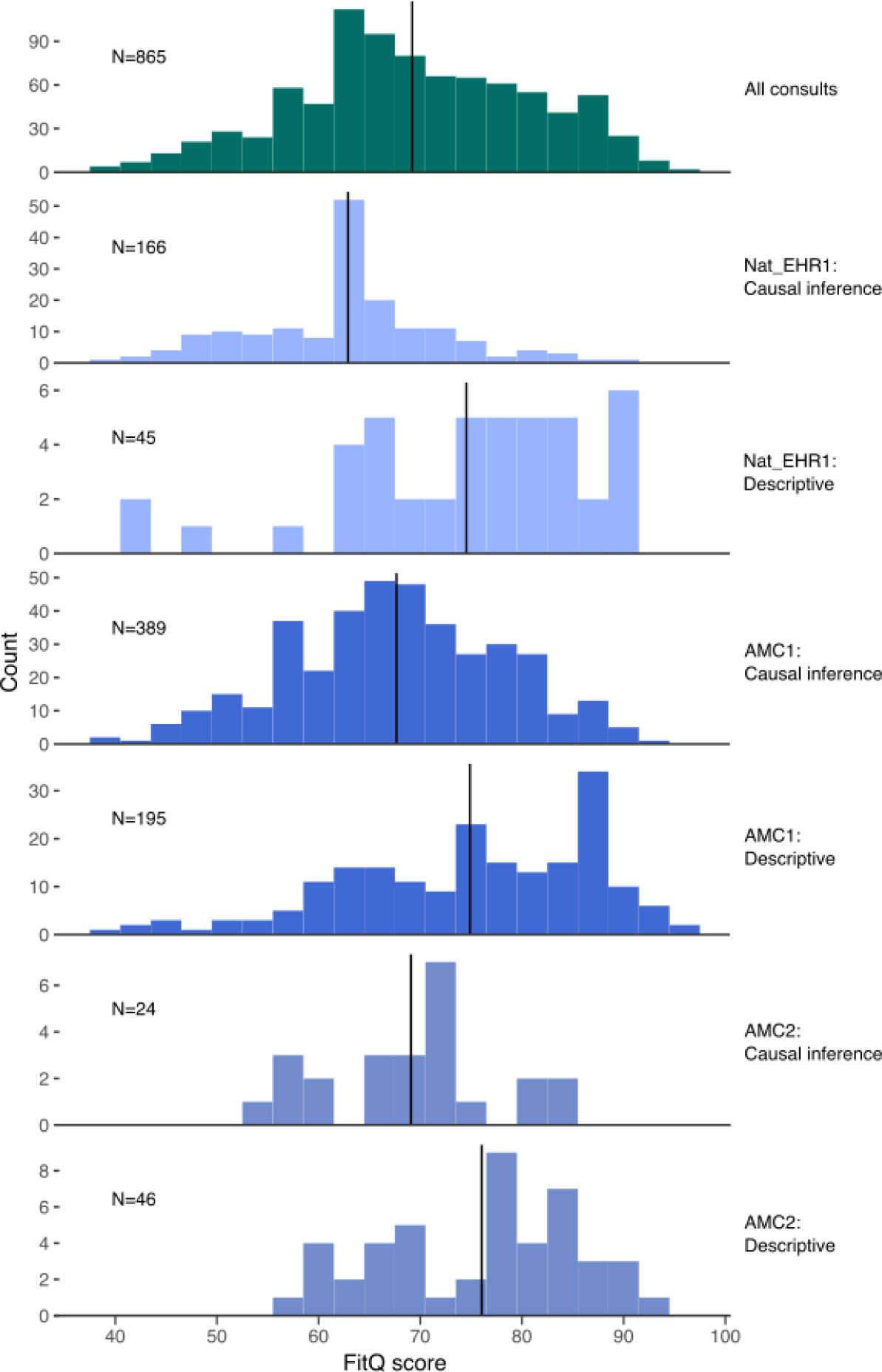
Distribution of FitQ scores for 865 RWE consultations, overall and by data source and purpose. Vertical bars indicate group-specific means.

Categorizing the consultations by whether they asked descriptive questions (289 consultations) or causal inference questions (576 consultations), FitQ scores tended to be higher for descriptive (mean 75.0, sd 11.8) than for causal inference consultations (mean 66.3, sd 10.3). This is largely due to descriptive consultations scoring higher than causal inference consultations on the metrics for cohort size (mean 6.4 vs. 5.8) and number of outcomes (mean 7.1 vs. 6.0).

One finding to note is that data sources with low overall RWDS (such as Nat_EHR1) can still be appropriate for answering some types of questions, as seen by the fact that Nat_EHR1 has an overall RWDS of 33 but can have FitQ scores of 80 or higher for certain queries. As we would expect, running the same RWE query on different data sources can yield different FitQ scores on each source (Figure 3). Importantly, we see that even though Nat_EHR1 has a lower RWDS than AMC1, Nat_EHR1 is better fit for some RWE queries than AMC1. Of 30 queries run on both data sources, 15 had a higher FitQ on Nat_EHR1, 14 had a higher FitQ on AMC1, and one had the same FitQ score on each data source.

**Figure 3:**
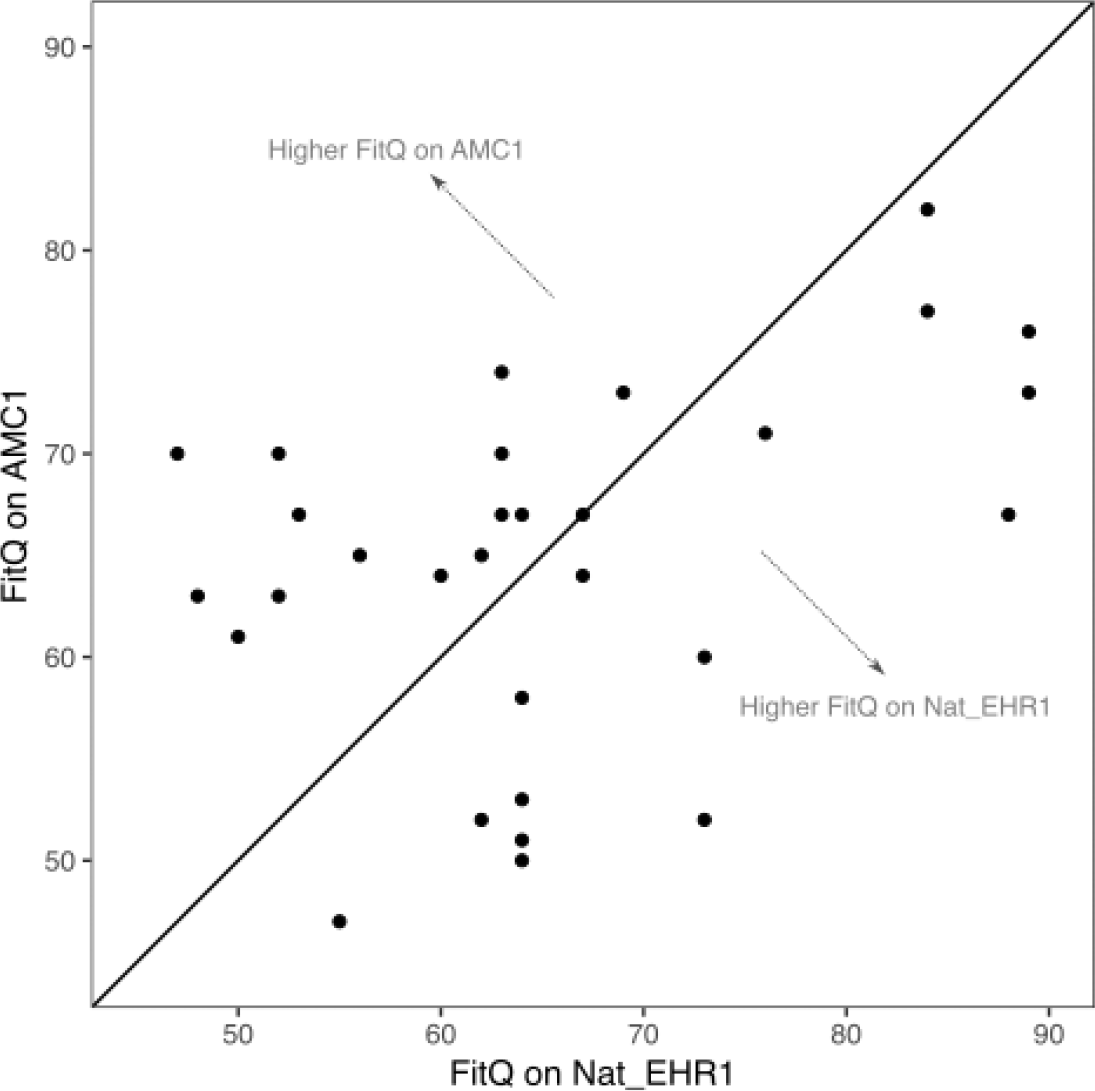
FitQ scores on Nat_EHR1 and AMC1 data sources when running the same query on each. The diagonal line represents queries with the same FitQ score on each data source.

We also find that RWDS elements can be predictive of the eventual FitQ scores, especially if certain diseases are underrepresented in the data set. In the RWDS, Nat_EHR1 scores low on the breadth metrics of patients with dermatological disease codes (0.06) and patients with nervous system diseases (0.08). Corresponding to this, the Nat_EHR1 FitQ scores in dermatology consults (63.8) and neurology consults (mean FitQ, 59.6) score below the Nat_EHR1 average FitQ of 65.4.

## Discussion

RWDS and FitQ fill a gap in the published literature, providing quantitative metrics of fitness-for-purpose of RWD sources for RWE questions. These scoring systems provide a flexible structure, where pre-computed quantitative metrics can be weighted either based on a standard approach or with use-case-specific weights. This approach allows users to quickly characterize the overall quality of RWD sources for general research purposes as well as the fitness of a specific RWD source for a specific RWE question. At Atropos Health, we currently use RWDS and FitQ to guide users of our portal to appropriate data sources and to assist with interpretation of query results. Applied more broadly, RWDS and FitQ can guide researchers in decisions about purchasing access to expensive RWD sources.

RWDS and FitQ also have applications beyond the traditional goal of identifying the best-fit data source for a single RWE study. The rise of networks linking many RWD sources from multiple providers is creating new paradigms for healthcare research and drug safety surveillance. Beyond traditional observational studies^18^ and pragmatic clinical trials^19^, these networks are empowering new uses of RWD. An example is OHDSI’s HowOften project, which aims to estimate the incidence rate of every potential medication side effect for every medication across all eligible OHDSI RWD sources.^20^ In these settings, the number of data sources, exposures, and outcomes of interest mean that evaluations of RWD fitness-for-purpose must be automated and run at scale. Our RWDS and FitQ scores provide a method for this automation. By pre-defining quantitative metrics of interest based on data or metadata from RWD sources, both these scores can be rapidly computed for any RWE question or comparison of interest.

An additional advantage of RWDS and FitQ is transparency. The metrics and weights used to calculate each score can be available along with the final scores themselves. This makes the data grading process clear and reproducible, which are particular requirements for RWE that may be used for regulatory or coverage decisions.^1,21^ A further use is to evaluate potential data drift in RWD sources that are periodically refreshed. By computing RWDS at each update, potential data quality issues can be surfaced quickly. Extensions to the RWDS or FitQ frameworks are also possible. For example, binary indicators such as presence/absence of claims data or cause of death data could be added as needed for specific use cases.

The primary limitation of RWDS and FitQ is the somewhat arbitrary nature of the chosen metrics and the weights or threshold values. Ultimately, any data grading system will require subjective decisions about metrics and cutpoints, but the values we have chosen may be less appropriate for other settings or systems. Second, while customized weights make RWDS flexible for certain use cases, changing the weights from query to query makes longitudinal comparisons of scores difficult. Finally, while high or low FitQs are self explanatory, mid-level FitQs are not always interpretable alone, and may require looking at the individual metrics to understand what factors have played into the score.

RWD sources are proliferating in healthcare, and RWE is playing an increasingly important role in clinical practice and in regulatory decisions. Systematic, transparent methods for evaluating RWD fitness-for-purpose are essential for effective use of RWD and for trustworthy RWE. RWDS and FitQ provide tools for producers and consumers of RWE to assess the quality of the RWD underlying the RWE.

## Data Availability

All data produced in the present study are available upon reasonable request to the authors

